# Pre-vaccination immunotypes reveal weak and robust antibody responders to influenza vaccination

**DOI:** 10.1101/2023.08.01.23293495

**Authors:** Alper Cevirgel, Sudarshan A. Shetty, Martijn Vos, Nening M. Nanlohy, Lisa Beckers, Elske Bijvank, Nynke Rots, Josine van Beek, Anne-Marie Buisman, Debbie van Baarle

## Abstract

Effective vaccine-induced immune responses are particularly essential in older adults who face an increased risk of immunosenescence. However, the complexity and variability of the human immune system make predicting vaccine responsiveness challenging. To address this knowledge gap, our study aimed to characterize immune profiles that are predictive of vaccine responsiveness using “immunotypes” as an innovative approach. We analyzed an extensive set of innate and adaptive immune cell subsets in the whole blood of 307 individuals (aged 25-92) pre- and post-influenza vaccination which we associated with day 28 hemagglutination inhibition (HI) antibody titers. Building on our previous work that stratified individuals into nine immunotypes based on immune cell subsets, we identified two pre-vaccination immunotypes associated with weak and one showing robust day 28 antibody response. Notably, the weak responders demonstrated immune regulation (HLA-DR+ T-cells) and activation (CD38+ T-cells) signatures respectively, while the robust responders displayed a high naïve-to-memory T-cell ratio and percentage of non-classical monocytes. These specific signatures deepen our understanding of the relationship between the baseline of the immune system and its functional potential. This approach could enhance our ability to identify individuals at risk of immunosenescence. Our findings highlight the potential of pre-vaccination immunotypes as an innovative tool for informing personalized vaccination strategies and improving health outcomes, particularly for aging populations.

## INTRODUCTION

Vaccines are widely recognized as the most effective means of protecting individuals from infectious diseases. However, the variability in effectiveness of influenza vaccines among older adults, ranging from 17-53%, leaving a significant proportion of the population unprotected^1–6^. One potential reason for this variability could be the phenomenon called immunosenescence, an age-related decline in immune function^1, 7^. Consequently, there is a pressing need to identify biomarkers of immune function at baseline. These biomarkers could help recognize individuals at risk of developing impaired responses to vaccines or pathogens and inform personalized vaccine strategies or treatments^8^. To this end, a deeper understanding of vaccine responsiveness and potential predictive biomarkers in aging adults is crucial.

Despite the importance of identifying biomarkers that predict influenza vaccine responses, the number of studies on this topic is limited. Previous studies have described potential pre-vaccination biomarkers such as CD4+ T memory^9–11^, CD8+ T memory^10–12^ and B memory^9, 11^ cells. Although these subsets are potential predictors of influenza vaccine responses, they exhibit high inter-individual variation influenced by factors such as age and chronic viral infections like cytomegalovirus (CMV)^13^. Moreover, the immune system’s functionality emerges from a complex network of interactions among various components, not fully represented by individual immune cell subsets, therefore, making single immune subsets poor biomarkers of vaccine responsiveness^14–16^. With an intent to overcome these limitations, we hypothesized that a combination of immune subsets could provide a more comprehensive representation of the immune network and offers more insightful correlations with immune functionality, for which we use antibody responses to influenza vaccination as a proxy.

To test our hypothesis, we analyzed an extensive number of immune subsets (percentages and absolute numbers) and antibody responses to influenza by assessing hemagglutination inhibition assay (HI) titers pre- and post- influenza vaccination in 307 individuals (25-92 years old). Baseline individual immune subsets exhibited limited association with day 28 HI titers, whereas stratification of individuals into immunotypes, clusters of individuals that share similar immune cell subset networks irrespective of their age, using an unsupervised approach based on 59 immune subsets did reveal associations with influenza antibody responses^13^. We identified immunotypes associated with weak or robust antibody responses and biologically interpreted these response patterns. Our findings suggest that pre-vaccination immunotypes associate with antibody responses to influenza vaccination and contain signatures that improve our understanding of immune aging and post-vaccination immune subset kinetics. This research could accelerate the translation of knowledge from our fundamental understanding of immunotypes to applications in personalized vaccination strategies and thereby maycontribute to the development of interventions that better protect aging populations.

## RESULTS

### Characteristics of the study population

To identify potential biomarkers of response to influenza vaccination, we recruited participants from the VITAL cohort study (N=308, Supp. Fig.1) aged 25 to 92 years old who received the quadrivalent inactivated influenza vaccine (QIV) in autumn 2019 (Fig. 1a)^17^. After vaccination, we tracked their cellular and humoral immune responses (Fig. 1b). We used previously reported data on immune cell subsets and measured hemagglutination inhibition (HI) titers against the influenza strains^13^. We focused on HI titers against the H3N2 (A/Kansas/14/2017) strain as other strains showed limited antibody responses (van der Heiden, et al. manuscript in prep).

**Figure 1:**
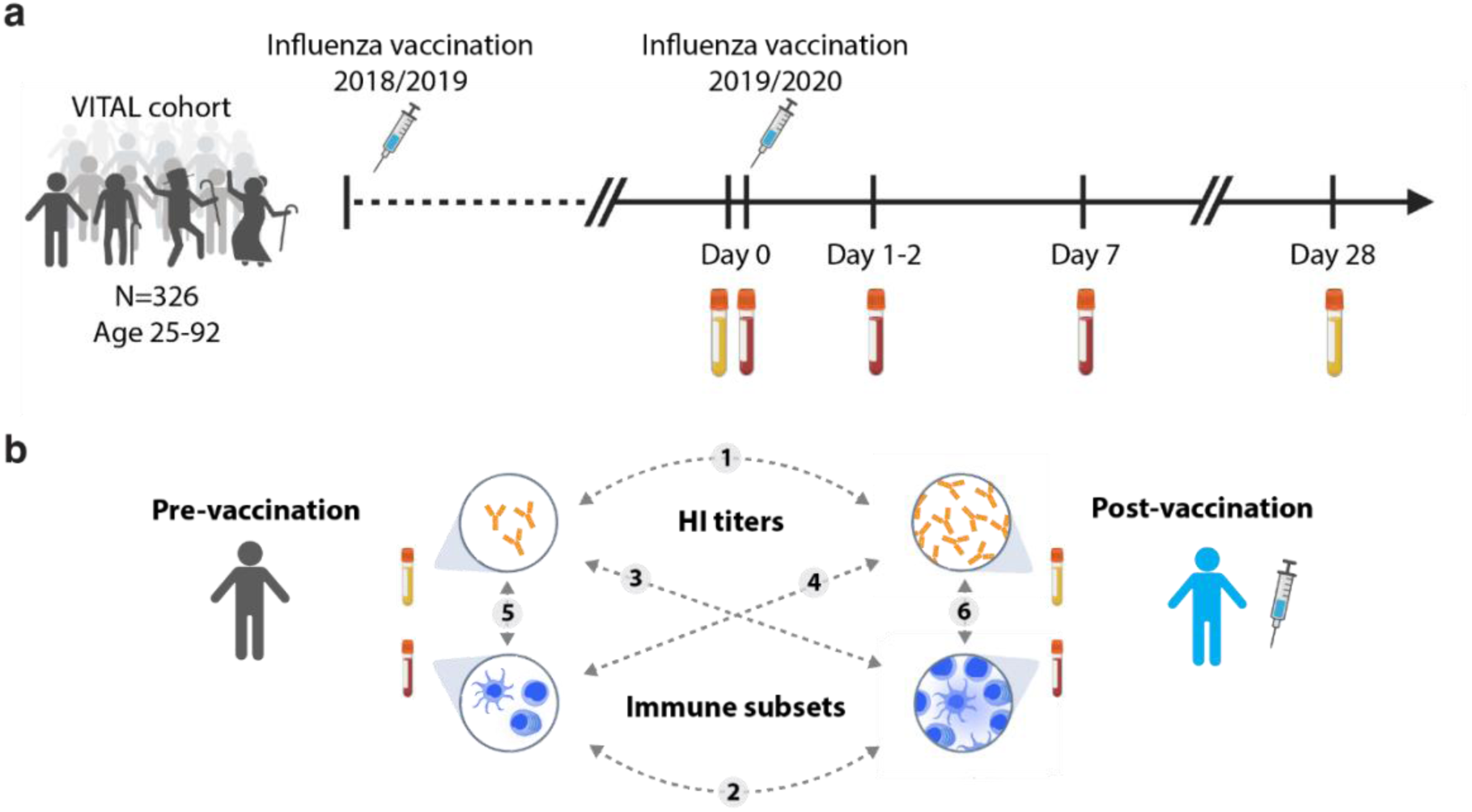
Clinical trial description. **(a)** Timepoints and sample collections**. (b)** Overview of relationships between pre-/post-vaccination hemagglutination inhibition (HI) titers and immune subsets investigated in this study. Dashed lines with numbers describe the relationships investigated in the study.

### Impact of pre-vaccination influenza antibodies on defining sero-responders

To investigate antibody responses to influenza virus, we first used the classical sero-responder definition (HI titer ≥ 40 and a four-fold increase in HI titer at day 28 compared to baseline). Out of 307 individuals, 190 (62%) were characterized as sero-responders. Before vaccination, an HI titer below the sero-protection threshold of 40 was observed in 207 out of 307 individuals, which constituted 67% of the total study group (Fig. 2a)^18^. At 28 days after vaccination, only 42 out of 307 (13.6%) participants had an HI titer below 40. (Fig. 2a).

**Figure 2:**
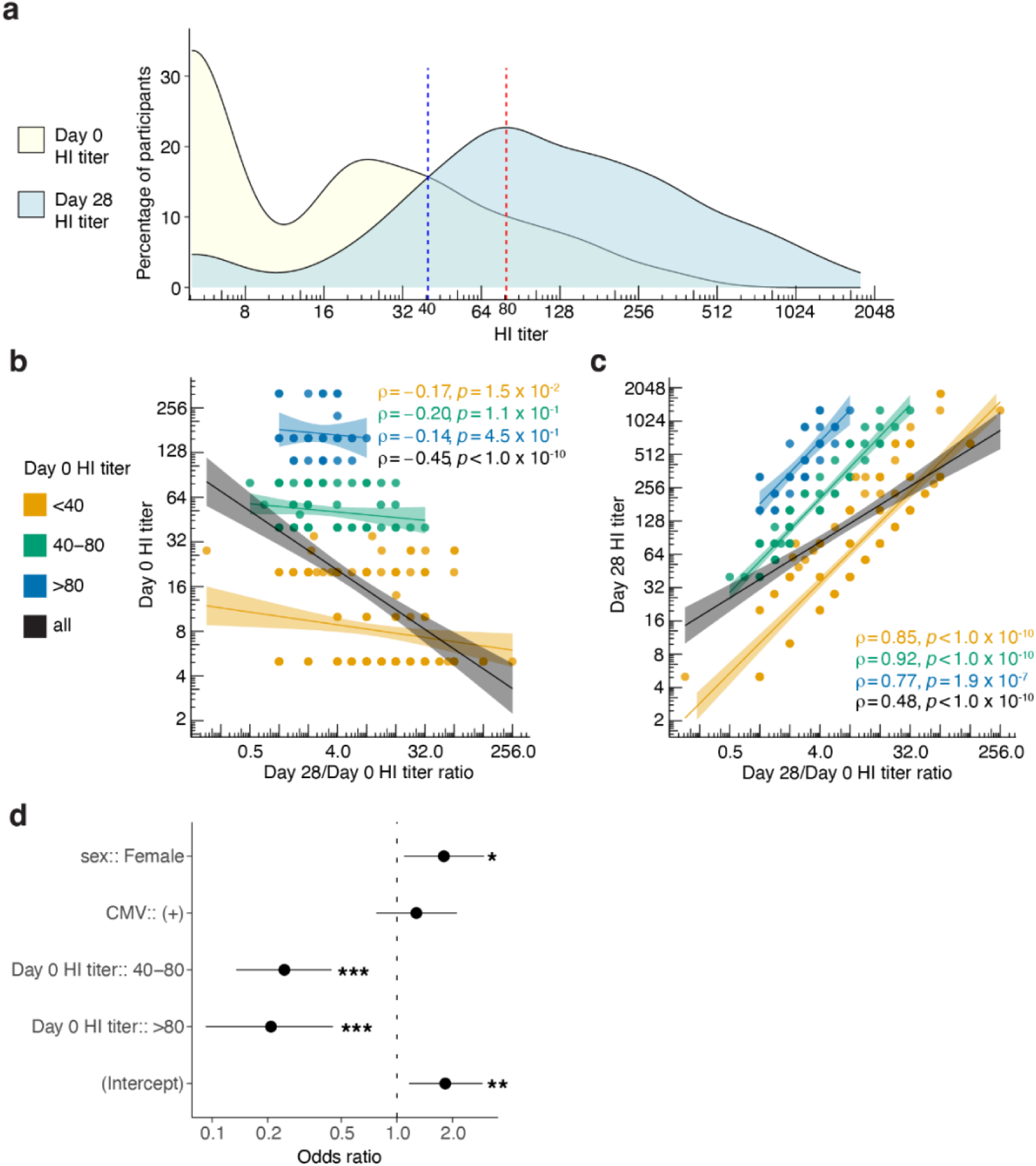
Influence of pre-vaccination HI titers on post-vaccination HI titer fold change and sero-responder categorization. **(a)** Distribution of participants based on HI titers at day 0 and day 28. Blue dotted line indicates sero-protection threshold of 40 and red dotted line indicates median post vaccination HI titer. **(b)** Spearman correlation between day 0 HI titers and day 28/day 0 HI titer ratio. **(c)** Spearman correlation between day 28 HI titers and day 28/day 0 HI titer ratio. Correlation coefficient rho (ρ) and p values are reported for each pre- vaccination HI titer group. **(d)** A logistic regression model demonstrating the odds of becoming a sero-responder to the influenza vaccination based on sex, CMV and pre-vaccination HI titers.

Pre-vaccination HI titers could influence the categorization of individuals as sero-responders since these have been previously described to correlate with post-vaccination HI titers (Fig. 1b-circle-1)^19^. We, therefore, stratified individuals into three groups based on their pre-vaccination HI titers, taking into account both the sero-protection threshold of 40 and the median post-vaccination HI titer of 80. This categorization resulted in groups representing sero-negative individuals with HI titers <40, low-sero-positive individuals with HI titers from 40-80, and high-sero-positive individuals with HI titers >80. In our data, indeed, pre-vaccination HI titers were negatively correlated to day 28/ day 0 HI titer fold change (ρ=-0.4, p<1.0x10^-10^). Next, we investigated whether pre-vaccination HI titer groups could reduce the influence of pre-vaccination titers on the HI titer fold change. We observed that in pre-vaccination HI titer groups, the correlation between pre-vaccination HI titers and HI titer fold change was improved (Fig. 2b). Additionally, within the HI titer groups, the correlation between day 28 HI titers and HI titer fold change was stronger compared to the correlation for the whole cohort (ρ=0.48) since the influence of pre-vaccination titers was minimized (Fig. 2c).

To further dissect other factors that influence the odds of being categorized as a sero-responder, we performed a logistic regression analysis to assess the influence of sex, cytomegalovirus (CMV) seropositivity and pre-vaccination HI titer groups. The odds of being a sero-responder in females were 80% higher (p=2.6x10^-2^, CI[1.18, 2.96]) than in males. CMV-seropositivity did not show a significant effect (p=3.8x10^-1^). Individuals in pre-vaccination HI titers of “40-80” and “>80” showed 75% (p=3.3x10^-6^, CI[0.14, 0.44]) and 81% (p=6.1x10^-5^, CI[0.08, 0.42]) lower odds of being categorized as sero-responder, respectively compared to “<40” (Fig. 2d). Thus, pre-vaccination HI titers should be taken into account to study antibody responses to influenza vaccine. Therefore, we integrated the pre-vaccination HI titers groups in our further downstream analyses on vaccine responsiveness to influenza.

### Post-influenza vaccination immune cell subset kinetics

Humoral responses to vaccination are orchestrated by specific B and T cell subsets in secondary lymphoid organs^20^. Thus far, post-vaccination changes in numbers of circulating follicular (CXCR5+) CD4+ T cells and plasmablasts (CD19+CD27+CD38+) at day 7 were described as some of the best correlates of antibody responses to influenza vaccination ^20, 21^.

Our analyses focused on which immune cell subsets (percentage and absolute number) before (day 0) and after (days 1-2, day 7) vaccination experienced the most significant changes (Dunn’s test). Increased cell numbers and percentages of follicular (CXCR5+) CD4+ T naïve (Tn)(CD27+CD45RO-) cells and non-classical monocytes (CD14dimCD16+) and decreased numbers and percentages of CD56dim CD38+HLA-DR+ Natural killer (NK) cells were observed as early as 1-2 days post-vaccination (Fig. 3a-c, Table 1). At 7 days post-vaccination, the increase in follicular CD4+ Tn cells was further amplified (Fig. 3a, Table 1), and additionally, significantly higher numbers and percentages of both plasmablasts and activated follicular (CXCR5+CD38+) CD4+ T cells were identified (Fig. 3d-e, Supp. table 1).

**Figure 3:**
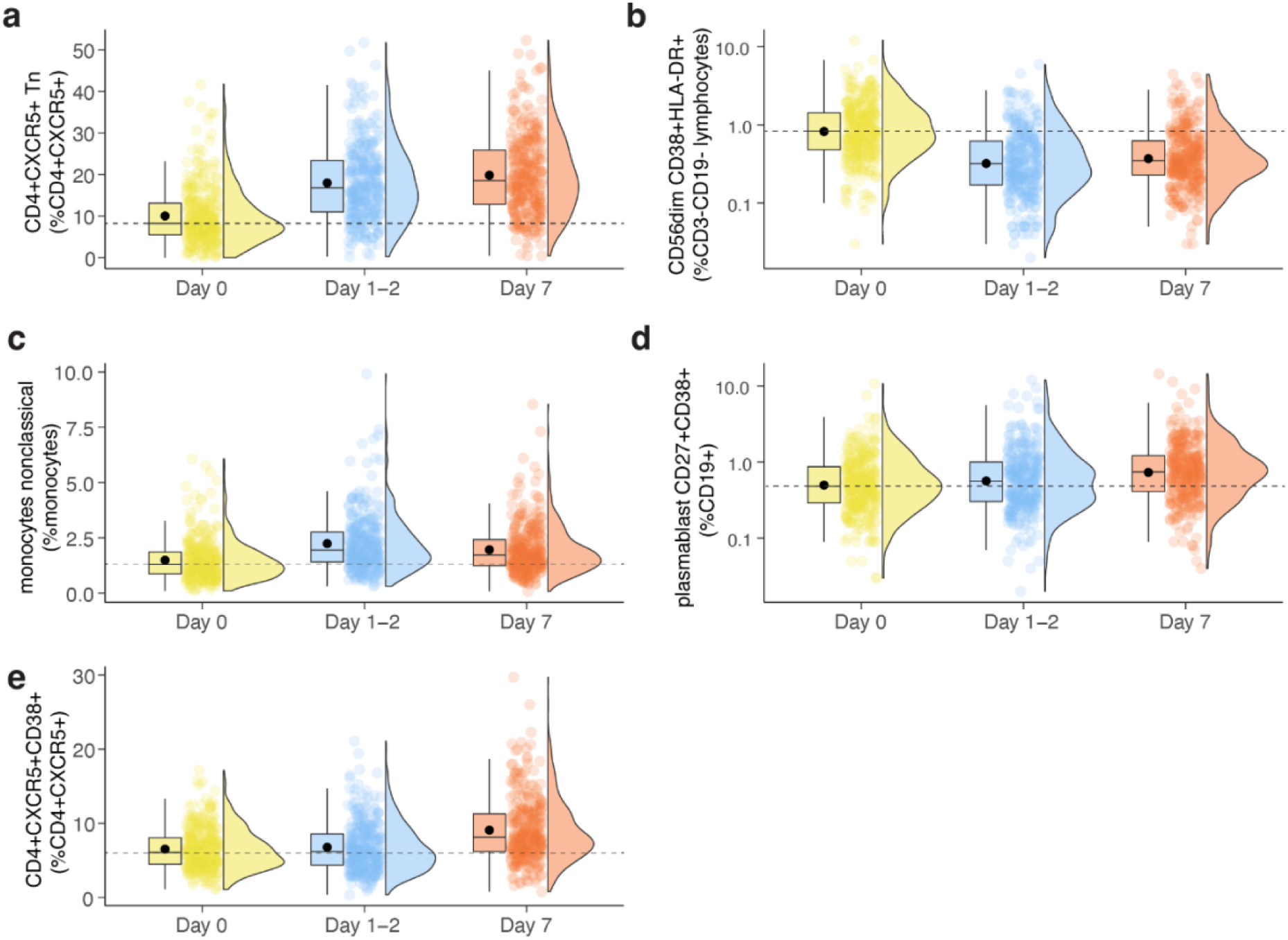
Pre- and post- influenza vaccination immune subset kinetics with the highest statistical significance. Boxplots depicting mean (dot) and median (line) day 0, day 1-2, and day 7 percentages of **(a)** follicular (CXCR5+) CD4+ T naïve (Tn, CD27+CD45RO-), **(b)** CD56dimCD38+HLA-DR+ Natural killer (NK) **(c)** non-classical monocytes (CD14dimCD16+) **(d)** plasmablasts (CD19+CD27+CD38+) and **(e)** activated (CD38+) follicular (CXCR5+) CD4+ T cells. Dashed line indicates the median value of day 0 for the given immune subset.

**Table 1:** Immune subsets with the highest significant changes at 1-2 days and 7 days post- vaccination. Percentages and absolute numbers of top 10 immune subsets are shown.

| Day 1-2/Day 0 immune subsets |  |  |  |  |  |  |  |
| --- | --- | --- | --- | --- | --- | --- | --- |
| variable (percentage) |  | p.adj | fc | variable (absolute number) |  | p.adj | fc |
|  | CD56dim CD38+HLA-DR+ | 3.94416E-31 | 0.40 |  | CD56dim CD38+HLA-DR+ | 9.28937E-31 | 0.41 |
|  | CD4+ CXCR5+ Tn | 5.36118E-31 | 1.92 |  | monocytes nonclassical | 1.66234E-24 | 1.64 |
|  | CD4+ CXCR5+ Tcm | 2.30582E-23 | 0.88 |  | CD4+ CXCR5+ Tn | 1.1087E-23 | 1.97 |
|  | CD56dim CD95+HLA-DR- | 2.39897E-21 | 0.45 |  | monocytes nonclassical HLA-DR+ | 3.35792E-20 | 1.63 |
|  | monocytes nonclassical | 5.34936E-19 | 1.52 |  | CD56dim CD95+HLA-DR- | 5.54292E-16 | 0.46 |
|  | monocytes classical | 1.43581E-18 | 0.95 |  | CD8+ Tcm CD95+HLA-DR- | 5.19481E-15 | 0.66 |
|  | CD56dim CD95-HLA-DR- | 8.37833E-16 | 1.02 |  | CD56dim CD95+HLA-DR+ | 2.65594E-12 | 0.53 |
|  | CD56dim CD95+HLA-DR+ | 4.16043E-14 | 0.55 |  | CD8+ CXCR5+CD95+ | 4.9424E-12 | 0.55 |
|  | CD4+ CXCR5+CD95+ | 4.53831E-14 | 0.84 |  | CD8+ CD95+HLA-DR- | 8.07561E-12 | 0.69 |
|  | CD8+ CD95+HLA-DR- | 8.23721E-14 | 0.73 |  | CD8+ CD95+ | 2.44069E-11 | 0.70 |
| Day 7/Day 0 immune subsets |  |  |  |  |  |  |  |
| variable (percentage) |  | p.adj | fc | variable (absolute number) |  | p.adj | fc |
|  | CD4+ CXCR5+ Tn | 2.84189E-41 | 2.17 |  | CD4+ CXCR5+ Tn | 2.91996E-42 | 2.86 |
|  | CD4+ CXCR5+ Tcm | 3.7126E-31 | 0.87 |  | CD56dim CD38+HLA-DR+ | 4.25727E-29 | 0.45 |
|  | CD56dim CD38+HLA-DR+ | 3.09541E-29 | 0.47 |  | CD4+ CXCR5+CD38+ | 2.66323E-27 | 1.79 |
|  | CD56dim CD95+HLA-DR- | 2.47147E-20 | 0.49 |  | CD4+ CXCR5+CD38+HLA-DR- | 1.36465E-25 | 1.73 |
|  | CD4+ CXCR5+CD38-HLA-DR- | 4.66271E-19 | 0.97 |  | CD4+ CXCR5+HLA-DR+ | 2.0469E-21 | 2.15 |
|  | CD56dim CD95-HLA-DR- | 2.04788E-17 | 1.02 |  | CD4+ CXCR5+ Tem | 6.46852E-18 | 1.78 |
|  | CD4+ CXCR5+CD38+ | 1.53147E-16 | 1.33 |  | CD4+ CD95-HLA-DR+ | 3.9182E-17 | 1.69 |
|  | monocytes classical HLA-DR+ | 2.66911E-15 | 0.74 |  | CD56dim CD95+HLA-DR- | 6.38607E-17 | 0.49 |
|  | CD4+ Tcm CD95-HLA-DR+ | 1.20994E-14 | 1.62 |  | CD4+ CXCR5+CD38-HLA-DR+ | 4.3846E-16 | 1.99 |
|  | CD4+ CXCR5+CD95+ | 1.47547E-14 | 0.85 |  | CD56dim CD95+HLA-DR+ | 7.30719E-14 | 0.50 |
|  | adaptive immune subsets | Kruskal–Wallis test followed by Dunn's test is used.Benjamini–Hochberg adjusted p-values (p.adj) and fold changes (fc) are reported. |  |  |  |  |  |
|  | innate immune subsets |  |  |  |  |  |  |

Considering the correlation between pre-vaccination antibody titers and post-vaccination antibody fold change, we determined whether the changes in immune subset numbers and percentages were also associated with pre-vaccination values. The correlations between the pre- and post-vaccination immune subsets revealed negative correlation coefficients ranging from -0.2 to -0.8 with a mean of -0.4 (Fig. 1b-circle-2, Supp. Table 2). Interestingly, these post-vaccination changes in immune subsets appeared to be independent of pre-vaccination HI titer groups (linear regression, adj.p>0.05) (Fig. 1b-circle-3, Supp. Table 3).

### Weak and robust vaccine responder profiles revealed by immunotypes

We conducted regression analyses to identify pre-vaccination immune cell subsets that may be associated with the day 28 influenza antibody response (Fig. 1b-circle-4). We factored in potential confounding variables such as sex, CMV-seropositivity, and pre-vaccination HI titer groups into our models.

At baseline, percentages of monocytes, IgD+CD27+CD95+ memory B cells and CD8+ T effector memory cells (Tem) were positively associated with day 28 HI titers, whereas, numbers of follicular HLA-DR+CD4+ and follicular CD38+CD4+ T-cells showed a negative association (p<0.05). Nevertheless, these correlations were marked by low coefficients and not significant after multiple test corrections (Benjamini-Hochberg, p.adj>0.05), indicating weak associations with antibody responses (Supp. Table 4).

We hypothesized that the combination of immune subset variables representing different aspects of the immune network would associate better with vaccine responsiveness than individual immune subset variables. Previously we had stratified individuals from the same cohort into 9 distinct immunotypes, in an unsupervised fashion, based on resemblances among 59 immune cell subsets, including T cells, B cells, NK cells, monocytes, and granulocytes^13^. In our current analysis, we examined how these previously identified immunotypes associate with pre-/post-vaccination HI titers and sero-responder profiles (Fig. 1b-circle-5). Pre-vaccination HI titers remained similar across immunotypes (Kruskal-Wallis, p=0.55) (Fig. 4a). At day 28, immunotype-2 showed the lowest HI titers. In contrast, immunotype-6 showed the highest -HI titers at day 28 and the highest day 28/day0 fold change in HI titer compared to other immunotypes (Fig. 4b-c). Additionally, after sub-categorizing sero-responders based on day 28 HI titers of 40-80 or >80 as low-sero-responder and high-sero-responder, respectively, individuals with immunotype-6 showed the highest whereas immunotype-2 showed the lowest percentage of high-sero-responders, although not statistically significant (chi-squared test, high-sero-responders, p=4.0x10^-1^) (Fig. 4d).

**Figure 4:**
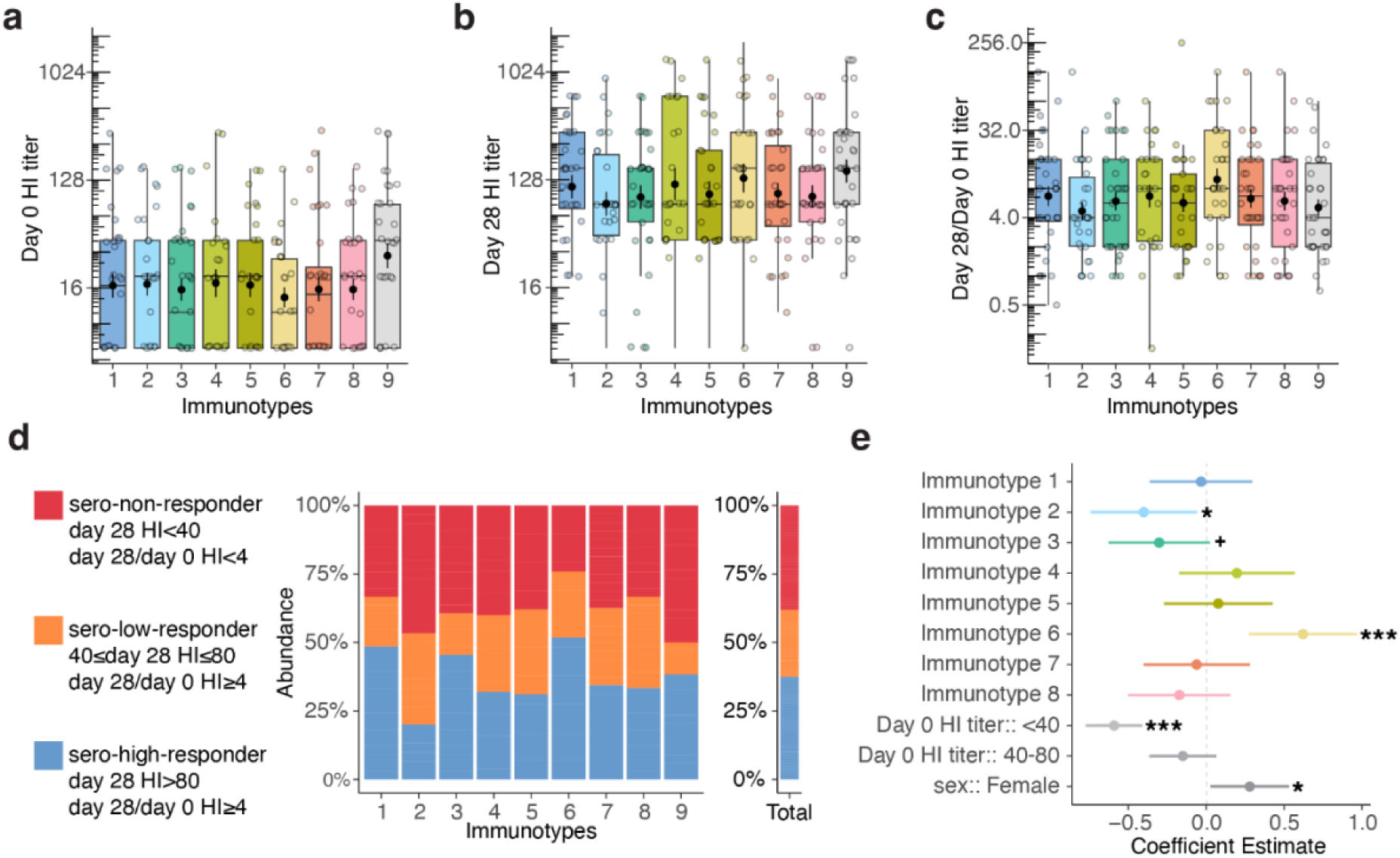
Identification of weak and robust vaccine responders through immunotypes. **(a)** Day 0 HI titers, **(b)** Day 28 HI titers, and **(c)** Day 28/day 0 HI titer ratio in immunotypes. **(d)** Comparative distributions of non-, low-, and high-sero-responder groups within different immunotypes and across the total cohort. **(e)** Regression models of day 28 HI titers for immunotypes after correcting for pre-vaccination HI titers and sex.

To confirm our findings above, we performed regression analyses of the day 28 HI titer, using immunotypes while adjusting variables that were significant for sero-responder categorization, namely sex and pre-vaccination HI titer groups. Among the 9 immunotypes, immunotype-2 and immunotype-3 showed negative coefficients (p=2.2x10^-2^ and p=6.9x10^-2^, respectively), whereas immunotype-6 exhibited a significant positive coefficient (p=4.9x10^-4^) for day 28 HI titers (Fig. 4e). These analyses further highlight the variation in antibody responses to influenza vaccination among individuals with different pre-vaccination immunotypes.

### Baseline and post-vaccination differences between weak and robust responder immunotypes

To elucidate the immune characteristics of immunotypes associated with weak and robust antibody responses, we compared their pre- and post-vaccination immune subset composition. Immunotype-6, which is associated with a robust antibody response, exhibited significantly higher T naïve (Tn) to T memory (Tm) ratios for both CD4+ and CD8+ cells and non-classical monocytes at baseline, compared to the rest of the immunotypes (Fig. 5a). These T cell subset ratios were previously described as aging-related immune phenotype markers^22^ that were associated with biological age and chronological age, respectively. At baseline, immune subset composition of immunotype-2 (associated with a weak antibody response), was dominated by increased percentages of HLA-DR expressing CD8+, CD4+ and follicular CD4+ T cell subsets as previously reported^13^ (Fig. 5a). In contrast, at baseline, immunotype-3 (associated with a weak antibody response) showed significantly higher Tn/Tm CD4+ and CD8+ ratio and percentages of CD38+ CD4+ Tregs, CD38+ CD4+ and CXCR5+CD38+ CD4+ T cells compared to rest of the immunotypes (Fig. 5a) which suggested an activated immune environment. In addition, both immunotypes-2 and -3, compared to the rest of the immunotypes, shared a cellular composition characterized by lower percentages of non-classical monocytes and CD95-HLA-DR- B cells at baseline (Fig. 5a)

**Figure 5:**
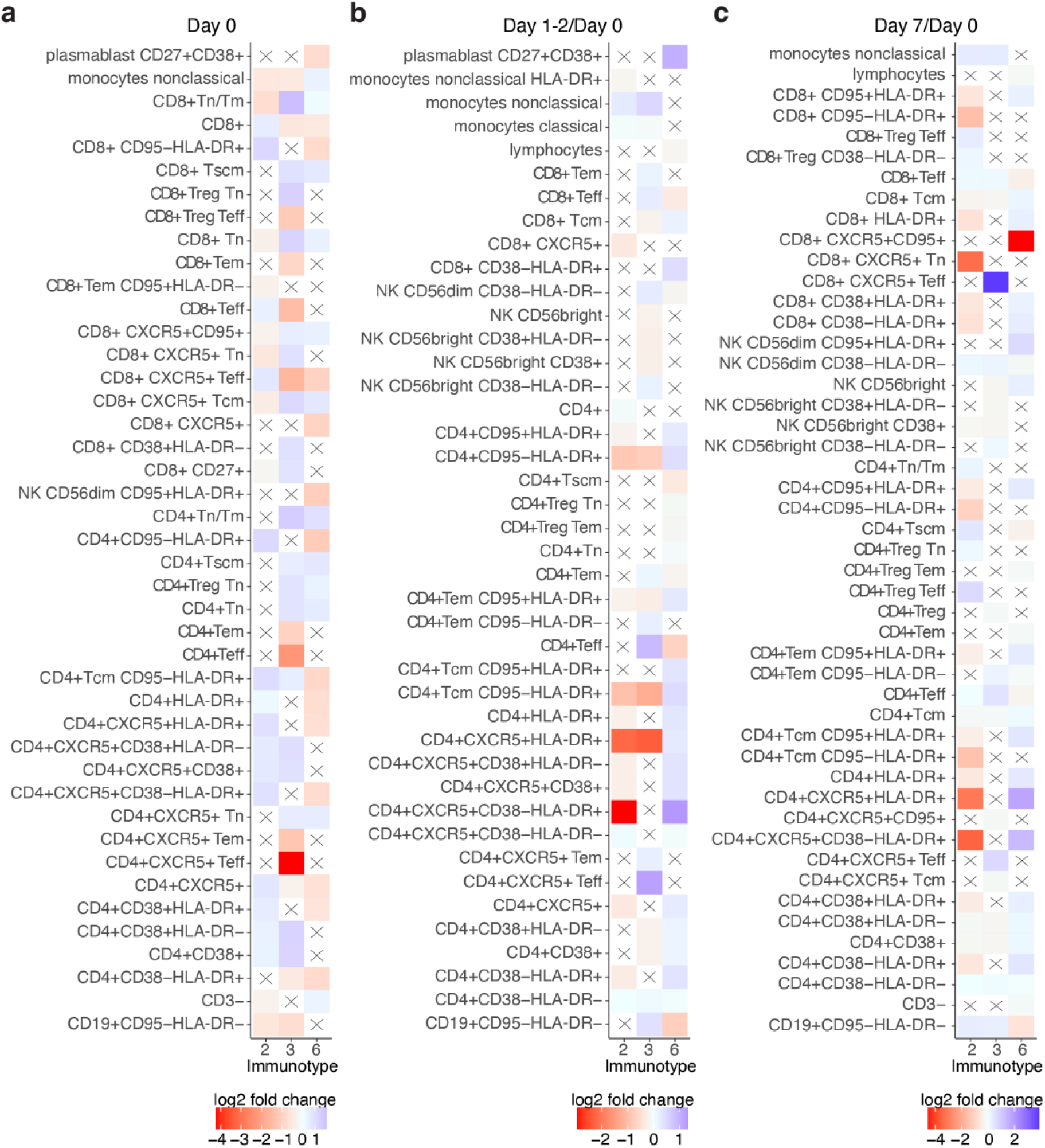
Pre- and post-vaccination cellular features of weak and robust responder immunotypes. Immune subset differences at **(a)** baseline, **(b)** 1-2 days and **(c)** 7 days after vaccination for weak responder immunotypes 2 and 3, and robust responder immunotype 6, each compared to the rest of the immunotypes. Only the top 10 immune subsets with the highest and the lowest fold changes, which are also statistically significant (adj.p≤0.05), are shown for each immunotype. An “X” symbol indicates a non-significant comparison.

At 1-2 days post-vaccination, the most predominant immune subset change for individuals classified in immunotype-6 compared to rest of the immunotypes was a rapid increase in percentage of CD38-HLA-DR+ follicular CD4+ T cells (Fig. 5b). In contrast, both immunotype- 2 and -3 did not show a similar early increase in percentage of HLA-DR+ follicular CD4+ T cells compared to the rest of the immunotypes (Fig. 5b). Unexpectedly, as early as 1-2 days post-vaccination, we observed an increased percentage of plasmablasts for individuals in immunotype-6 (Fig. 4b).

At day 7, the most prominently increased immune subsets in immunotype-6 compared to the rest of the immunotypes were percentages of HLA-DR+ follicular CD4+ T cells and other HLA-DR+ T cell subsets (Fig. 5c). Percentages of activated follicular CD4+ T cells and plasmablasts were also significantly increased in immunotype-6 (Supp. Table 5). Both immunotype-2/3 showed a significant increase in percentages of T effector (Teff) cell subsets at day 7 compared to the rest of the immunotypes. For immunotype-3, the highest increase was observed in percentage of follicular CD8+ Teff cells, whereas for immunotype-2, CD4+ Teff Tregs showed the highest increase (Fig. 5c). Remarkably, in immunotype-6, percentage of CD95+ circulating follicular CD8+ T cells showed a significant decrease.

### Non-activated follicular CD4+ T cell increase exhibits superior post-vaccination association with influenza antibody responses

We showed that individual immune subset variables at pre-vaccination failed to associate with influenza antibody responses, whereas specific immunotypes did associate with weak or robust vaccine responders. Next, we investigated the associations between post-vaccination immune subset kinetics and day 28 HI titers in regression models (Fig. 1b-circle-6), while adjusting for factors such as sex, CMV-seropositivity and pre-vaccination HI titers (Supp. Table 4, Fig. 6a).

**Figure 6:**
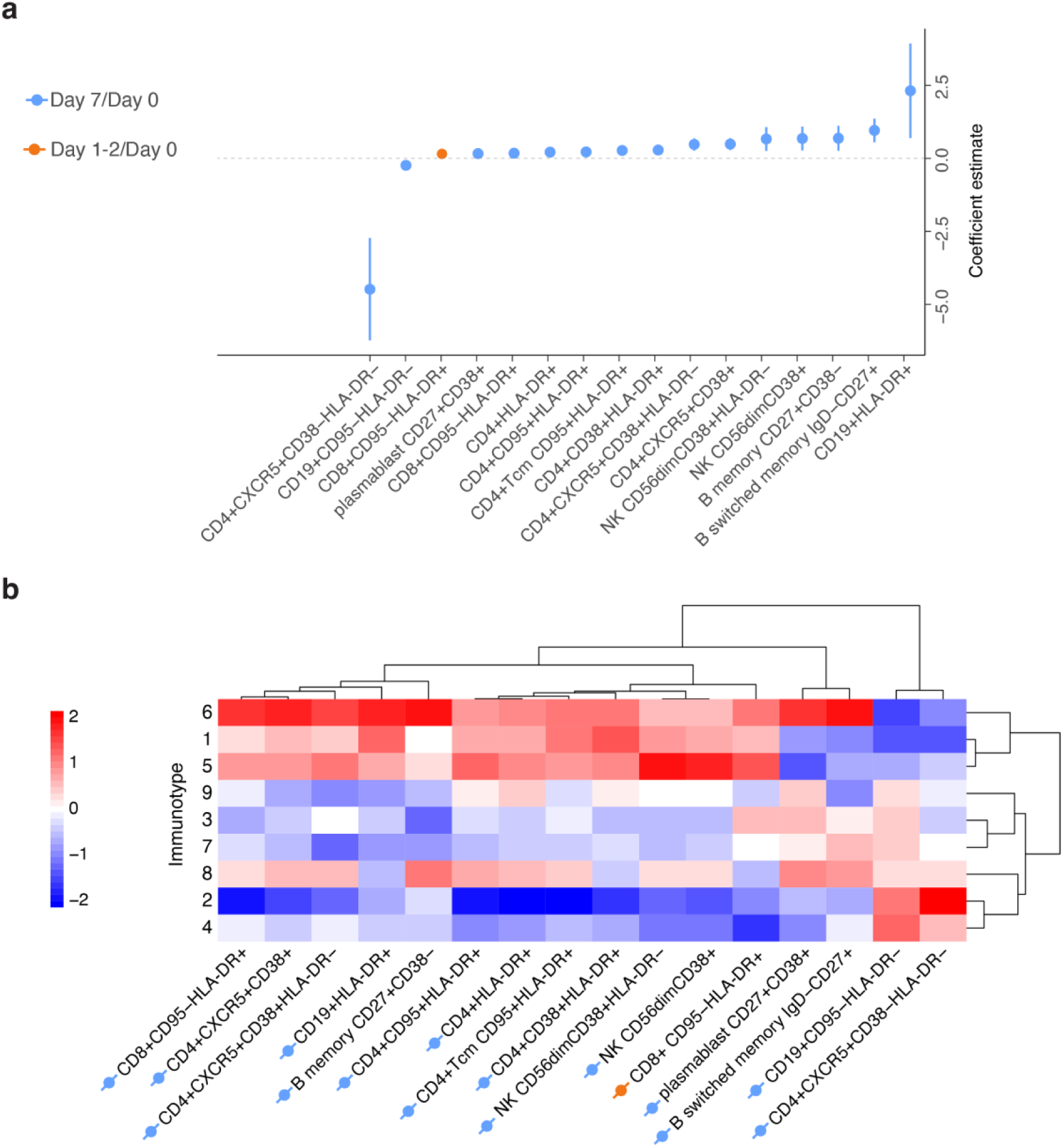
An increase in the percentage of non-activated follicular CD4+ T cells at day 7 is the strongest associate of antibody response. **(a)** Regression models of day 28 HI titers using pre-vaccination immune subsets and post-vaccination immune subsets kinetics, corrected for sex, CMV-seropositivity, and pre-vaccination HI titers. Percentages of the immune subset that are significant after false discovery rate correction are shown. Blue and orange indicate fold changes of day 7/day 0 and day 1-2/day 0 immune subsets, respectively. **(b)** A heatmap of post-vaccination fold changes in immune subsets significantly associated with antibody response is shown for each immunotype. Dendrograms are calculated based on correlation distance.

An increase in percentage of non-activated (CD38-HLA-DR-) follicular (CXCR5+) CD4+ T cells at day 7 compared to day 0 showed a stronger but negative association (larger absolute coefficient value) with antibody responses (adj.p= 1.1x10^-4^) than activated (CD38+) follicular CD4+ T cell (adj.p= 2.0x10^-4^) and plasmablasts (adj.p=1.4x10^-2^) (Fig. 6a). To validate our findings, we independently applied Ridge and random forest regression analyses to the same dataset. In both models, an increase in percentage of non-activated follicular CD4+ T cells demonstrated the strongest variable importance to post-vaccination antibody responses (Supp. Fig.2a-b).

Next, we hypothesized that the increase or decrease of specific post-vaccination immune cell subsets we identified in our regression models would be linked to the responder profile of immunotypes. Indeed, immunotype-6, which is associated with robust antibody response, had higher fold changes of subsets that are positively associated with antibody responses. In contrast, immunotype-2 and -3, which are associated with weak antibody response, had decreased fold changes for those immune subsets (Fig. 6b). Thus, specific immunotypes not only associate with antibody responses but also larger increases in cell subsets associated with antibody responses.

## DISCUSSION

Improving vaccine effectiveness is crucial, especially considering older adults who may be more susceptible to immune dysfunction and suboptimal vaccine responses. Identifying those at higher risk of immune dysfunction is of paramount importance, as this would enable us to devise personalized vaccination strategies and possibly implement alternative therapeutic approaches for their protection^8^. In this study, we aimed to determine immune subset profiles of at-risk individuals by examining the links between pre- and post-vaccination immune subsets and influenza antibody titers. We showed that pre-vaccination immunotypes, based on 59 immune subsets representing specific immune profiles, were associated with weak (immunotype-2 & 3) and robust (immunotype-6) responsiveness to influenza vaccination^13^.

Predicting post-vaccination humoral and cellular responses based on pre-vaccination numbers of immune cell subsets remains challenging for two main reasons. Firstly, substantial inter-individual immune variation makes it difficult to identify biomarkers^13, 23, 24^. Secondly, the relationship between pre-vaccination immune profiles and the functional capabilities of these immune profiles is yet to be fully explored. Although several studies investigated the associates of influenza vaccination using pre- and post-vaccination immune cell subsets, these studies focused on individual immune subsets as predictors of vaccination responses and did not explain the immune profiles of weak responders at pre-/post-vaccination^9, 10, 25^. We argue that such previously identified immune subsets provide incomplete insight into vaccine responsiveness since immune functionality is an emergent property of the intricate immune network, and hence, cannot be entirely attributed to individual immune cell subsets^14–16^. We hypothesized that a broader perspective, focusing on the composition of immune subsets, could provide a more holistic representation of the immune landscape and functionality than individual immune subsets. Our findings endorse this hypothesis as immunotype-2 (weak response-associated) showed immune signatures of aging-related immune subsets and a highly remodeled immune profile^13^. Moreover, immunotype-2 exhibited this perturbated immune network and lower immune stability despite being younger on average than immunotype-6 (robust response-associated), with a median age of 55 and 69 respectively^13^. This suggests that, aside from chronological age, the composition of the immune network also holds substantial significance in vaccine responsiveness.

The conventional sero-responder categorization based on day 0 and day 28 HI titers could result in overlooking individuals who may not meet the standard criteria for responders due to a negative correlation between pre-vaccination HI titers and day 28/ day 0 HI titer fold change^26^. This limitation further complicates the identification of at-risk individuals. Changes in post-vaccination immune subsets could be important additional variables to study impaired vaccine responses and aid identification of at-risk individuals. Because unlike day 28 HI titers, post-vaccination immune subset changes were not associated with pre-vaccination HI titers. We observed that HLA-DR+ follicular CD4+ T cells increase as early as 1-2 days after vaccination whereas CD38+ follicular CD4+ T cells peak 7 days after vaccination. Lack of HLA-DR and CD38 expression and accumulation of these non-activated (CD38- HLA-DR-) circulating follicular (CXCR5+) CD4+ T cells at day 7 were a superior associate of antibody response compared to previously known correlates such as CD38+ follicular CD4+ T cells and plasmablasts. We postulate that these non-activated follicular cells could be either “cell activation failures” or “actively suppressed.” Further in-depth phenotyping of non-activated follicular CD4+ T cells may reveal their heterogeneity and markers of inhibition/modulation which could help to explain these mechanisms.

After examining the alterations in immune subsets and their associated antibody responses following vaccination, we discovered that immunotypes 1, 5, and 6 were more similar to each other than the other immunotypes. This was determined by analyzing the correlation distances on a dendrogram. Notably, these immunotypes exhibited a similar relationship based on their pre-vaccination immune subsets which we reported previously^13^. This highlights the strong connection between pre- and post-vaccination immune networks. However, among the three closely related immunotypes only immunotype-6 demonstrated a statistically significant difference in achieving higher day 28 antibody titers in our models.

In the context of influenza vaccination, we showed that the pre-vaccination state of the immune system is associated with antibody responses. Another example of the importance of regulation/activation was observed in immunotype-3 (weak response-associated) which did not exhibit aging-related immune signatures, such as lower Tn/Tm ratio, but an activated immune landscape at pre-vaccination, characterized by the percentage of CD38+ cells in total CD4+ T cells, follicular CD4+ and regulatory CD4+ T cell subsets. An activated immune environment with increased activated regulatory cells could hamper the development of potent immune responses which may explain the weaker response we observed in immunotype-3^27^.

Our study is not without limitations. While immunotypes-2/-3-6 display significant differences in antibody responses, there remains considerable variation in their antibody responses within these phenotypes. Since immunotypes contain individuals from various age groups these individuals are likely to have encountered different influenza strains throughout their lives^13^. This leads to a phenomenon known as immune imprinting or original antigenic sin where immune memory of a pathogen’s initial strain could limit immune system’s ability to respond to a new strain could be a factor in this variation^28^. Moreover, our findings on influenza vaccination may not translate into other types of vaccines or cohorts. Future research should investigate whether baseline immune profiles associate with vaccine responsiveness in different older populations and vaccine platforms.

In conclusion, our study emphasizes that the composition of pre-vaccination immune subsets, or “immunotypes,” may serve as a superior indicator of the functional capacity of the immune response to vaccination, as compared to individual immune cell subsets. This approach could potentially identify individuals at risk of suboptimal vaccine response, thereby guiding the development of more targeted and personalized vaccination strategies. In a broader context, these insights hold promise not only for evaluating vaccine responsiveness but also for understanding immune function in general, which could pave the way to a more profound comprehension of immune health.

## MATERIALS AND METHODS

### Cohort description and sampling

The Vaccines and InfecTious diseases in the Aging popuLation (VITAL) is a cohort started in 2019 in the Netherlands as described previously^13, 17^. In short, VITAL contains young, middle-aged and older adults (N=326) (aged 25 to 98 years old) who were vaccinated with the previous year’s seasonal influenza vaccine in 2018-2019. In 2019-2020, these individuals were vaccinated with the seasonal quadrivalent inactivated subunit influenza vaccine (QIV); Influvac Tetra (Abbott Biologicals B.V. The Netherlands) which contained neuraminidase and hemagglutinin from the following viral strains: A/Brisbane/02/2018, IVR-190(H1N1); A/Kansas/14/2017, NYMC X-327 (H3N2); B/Maryland/15/2016, NYMC BX-69A (B/Victoria/2/87 lineage); and B/Phuket/3073, wildtype (B/Yamagata/16/88 lineage) (Abbott Biologicals B.V. The Netherlands). For immune phenotyping whole blood samples (day 0, day 1-2, day 7) and for hemagglutination inhibition assay (HI) serum samples (day 0, day 28) were collected (Fig. 1a).

### Flow cytometry immune phenotyping

Flow cytometry and immune subset gating was performed as recently described^13^. In short, whole blood samples before (day 0), and after vaccination (day 1-2, day 7) were stained with anti-human fluorophore-conjugated antibodies and acquired on a 4-lasr LSRII Fortressa X20 flow cytometer (BD Biosciences). Absolute number of cells was calculated from event counts of TruCOUNT beads (BD Biosciences). Both percentage and absolute numbers of immune subsets were studied since the former describes the relative abundance of a subset in the parent population whereas the latter representsß the changes in total immune cell counts.

### Detection of baseline anti-influenza virus antibodies

To investigate antibody responses to QIV, antibodies against the H3N2 (A/Kansas/14/2017) strain were measured at Viroclinics (Rotterdam, the Netherlands) using the Hemagglutination Inhibition assay (HI) according to the standard methods of the World Health Organization (WHO) as explained in (refs from Josine: Rosendahl Huber; Frontiers Immunology 2019; Luytjes et al. Vaccine 2012; ECDC 2018). In short, a dilution series of serum samples was incubated with four Hemagglutinin Units (HAU) influenza virus for 20 minutes and thereafter incubated for 30 minutes with 0.25% turkey erythrocytes. Subsequently, the antibody titer (geometric mean titer) was determined as the reciprocal of the highest dilution of serum that prevents complete hemagglutination wells.

### Cytomegalovirus seropositivity

Immunoglobulin G antibodies against Cytomegalovirus (CMV) were quantified in serum by a multiplex immunoassay developed in-house and CMV-seropositivity thresholds were adapted from a previous study, as recently described^13^.

### Statistical modeling

Generalized linear model with binomial distribution (GLM) was built from sex, CMV-seropositivity and pre-vaccination HI titer groups to evaluate the effects on sero-responder categorization. The GLM’s Logit estimates were converted into Odds Ratio values through exponential transformation. For day 28 HI titer regression models, which is count data, negative binomial distribution showed a good fit to antibody data. Therefore, Generalized Additive Models for Location Scale and Shape (GAMLSS) function from “gamlss” R package which allowed negative binomial distribution were used^29^.

### Clustering analyses

At pre-vaccination, individuals were clustered into immunotypes as recently described^13^. In short, pairwise spearman correlation matrix of individuals based on 59 baseline immune subsets was calculated. The number of clusters (immunotypes) was decided based on Gap statistics and the data was clustered using k-means.

### Statistical analysis

Statistical analyses were performed using R (v4.2.2) and Rstudio (v2022.12.0+353). The sero- responder categorization based on pre-vaccination HI titers, sex, and CMV was implemented via the General Linear Models (GLM) from stats package (v4.2.2). Day 28 HI titer regression models were built using the GAMLSS package (v5.4.12). Heatmaps were generated with the pheatmap library (v1.0.12), and rstatix package (v0.7.2) was used for additional statistical analyses. For group comparisons of immune subsets at day 0, day 1-2, and day 7, the Kruskal– Wallis test was employed, followed by Dunn’s test when p-values were found to be significant. P values were adjusted for multiple comparisons using the Benjamini–Hochberg correction and reported as “adj.p”. Statistical significance is indicated in tables as follows: **^+^**p < 0.1 *p < 0.05, **p < 0.01, ***p < 0.001, ****p < 0.0001.

## List of Supplementary Materials

**Table S1:** Differences in immune subsets between days 0 and 1-2, and between days 0 and 7.

**Table S2:** Correlations between pre- and post-vaccination immune subsets.

**Table S3:** Regression models investigating the association of post-vaccination immune subset kinetics with pre-vaccination HI titer groups, sex, and CMV status.

**Table S4:** Regression models analyzing the influence of immune subsets (measured on days 0, 1-2, and 7) on day 28 antibody titers, accounting for pre-vaccination HI titer groups, sex, and CMV status

**Table S5:** Immune subset comparisons between immunotypes 2-3-6 and rest of the immunotypes for day 0, day 1-2 and day 7.

## Data Availability

Research data are not shared since the primary endpoints are not yet published.

## Acknowledgments

We thank Markus Viljanen for day 28 antibody response modeling, Marieke van Heiden for reading and giving feedback on the manuscript and Megan Barnes, Lysanne Bakker, Silvia Cohen, Shirley Man, Ilse Akkerman, Ruben Wiegmans for their help in handling the clinical samples. Human and vial icons in Figure 1 are created by BioRender.com.

## Funding

The VITAL project has received funding from the Innovative Medicines Initiative 2 Joint Undertaking (JU) under grant agreement No. 806776 and the Dutch Ministry of Health, Welfare and Sport. The JU receives support from the European Union’s Horizon 2020 research and innovation programme and EFPIA-members.

## Author contributions

AC, SAS, NR, AMB, and DvB conceptualized the study. AC, SAS, and NMN performed methodology. AC, SAS, MV, NMN, LB, JvB and NR performed the study. AC and SAS visualized the data. NR, AMB, and DvB were involved in funding acquisition. LB, EB, JvB, and DvB were involved in project administration. SAS, AMB, and DvB supervised the study. AC wrote the original draft. AC, SAS, AMB, DvB are involved in writing—reviewing and editing.

## Competing interests

Authors declare that they have no competing interests.

## Data and materials availability

Research data are not shared since the pre-clinical trial is ongoing and the primary endpoints are not yet published. The codes used in the manuscript are available from GitHub (https://github.com/alpercevirgel/Immunotype-Influenza-Response)

---

**Supplementary Figure 1:**
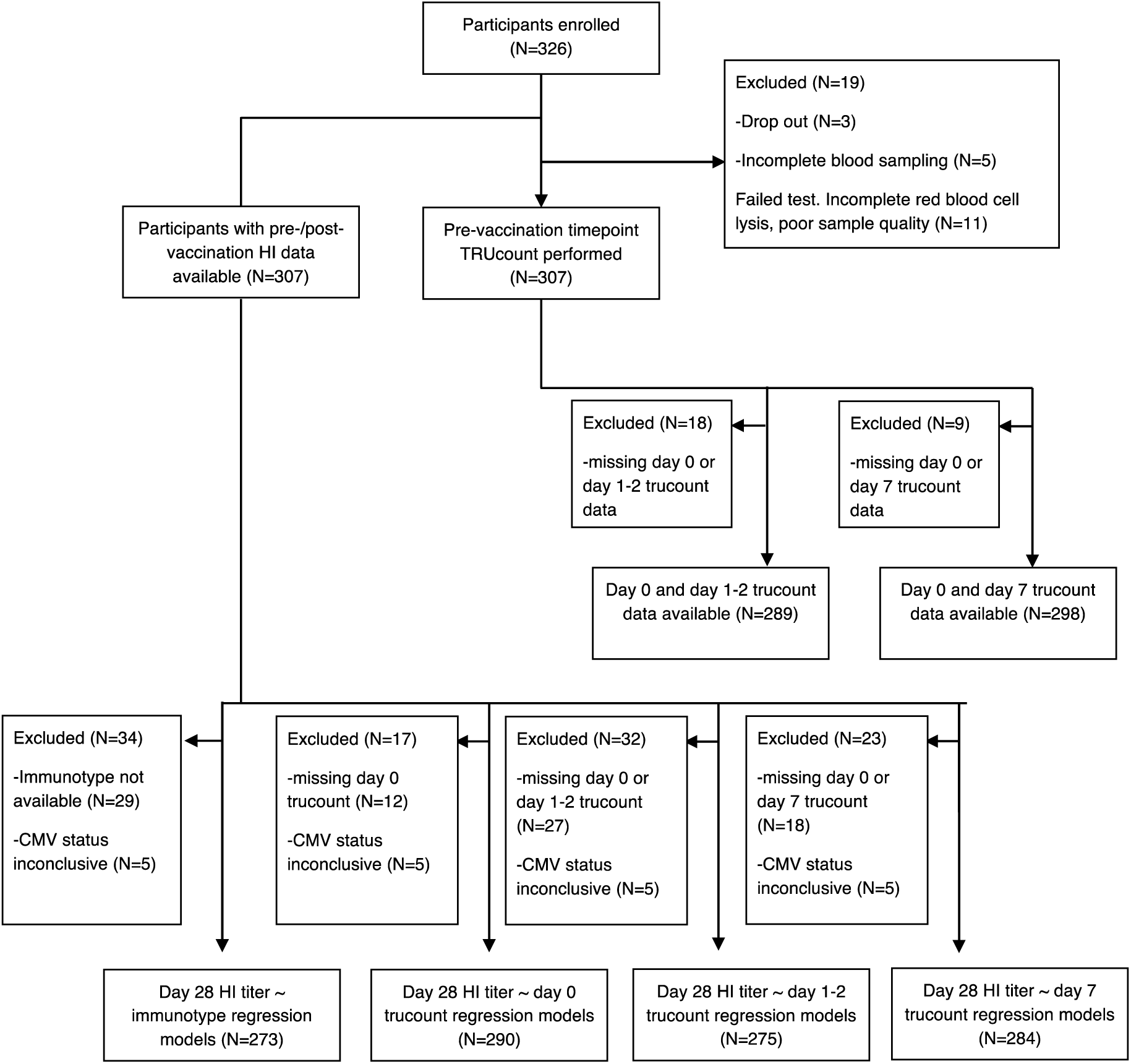
Flowchart of data analyses.

**Supplementary Figure 2:**
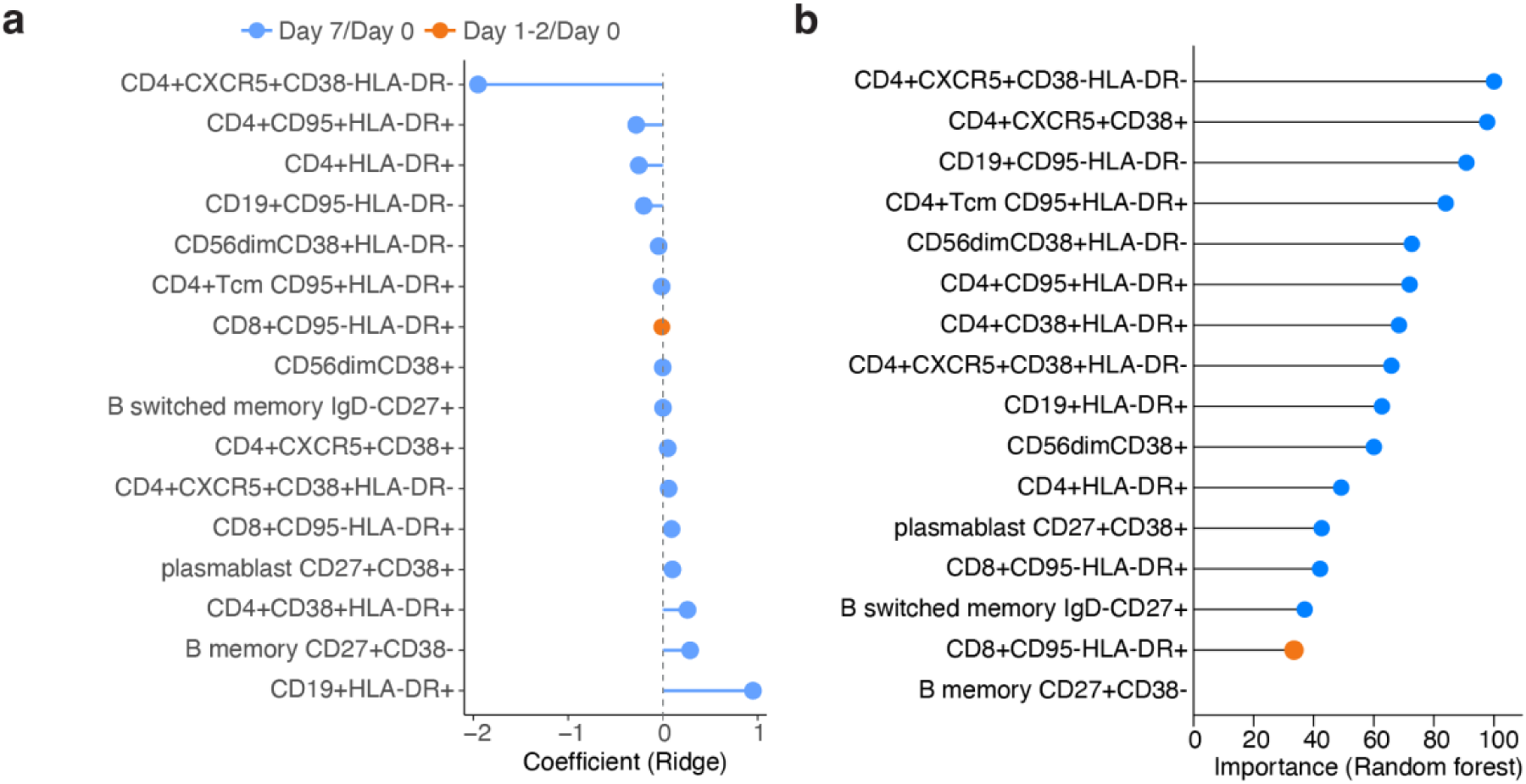
Percentage of non-activated follicular CD4+ T cell increase at day 7 is the most important associate of antibody responses in ridge & random forest regression models. (**a**) Coefficients (Ridge regression) and (**b**) variable importance (random forest regression) of day 28 HI titer regression analyses. Percentages of significant immune subsets (adj.p<0.05) that associated with antibody responses are used in both regression analyses.

## Notes

### Competing Interest Statement

The authors have declared no competing interest.

### Clinical Trial

EudraCT 2019-000836-24

### Author Declarations

Ethical approval was obtained through the Medical Research Ethics Committee Utrecht (NL69701.041.19, EudraCT: 2019-000836-24). All participants provided written informed consent and all procedures were performed with Good Clinical Practice and in accordance with the principles of the Declaration of Helsinki.

